# Machine Learning Applied to Clinical Laboratory Data Predicts Patient-Specific, Near-Term Relapse in Patients in Medication for Opioid Use Disorder Treatment

**DOI:** 10.1101/2020.08.10.20163881

**Authors:** Paul Pyzowski, Barbara Herbert, Wasim Q Malik

## Abstract

We have developed a data-driven, algorithmic method for identifying patients in an out-patient buprenorphine program at high risk for relapse in the following seven days. This method uses data already available in clinical laboratory data, can be made available in a timely matter, and is easily understandable and actionable by clinicians. Use of this method could significantly reduce the rate of relapse in addiction treatment programs by targeting interventions at those patients most at risk for near term relapse.

## 1 Introduction and Motivation

Addiction is a chronic disease, many patients will relapse during the course of treatment, and expectations of total abstinence from the first day of treatment are unrealistic for most patients. A patient may attend a medication for opioid use disorder (MOUD) treatment program ^(1)^involving buprenorphine or methadone for months or even years, with occasional relapses but generally on a path to recovery with less use and misuse of illicit substances, with commensurate improvements in quality of life.

Relapses come for different reasons - economic or social stressors, misuse of prescribed medications and/or illicit substances, and idiosyncratic cravings. In clinical practice relapses are difficult to predict. A patient may be using substances in a controlled, subthreshold manner that is not flagged on drug screening tests. There may be factors or events in a patient’s life that may not come to the attention of the clinician but act as stressors or triggers. Regardless of the reason, A clinician that had knowledge that a given patient was at high risk for relapse in the near term could make a targeted intervention or change in treatment (e.g. more frequent drug screening) could be made before the relapse occurred, with the goal of preventing a disruptive, expensive, and potentially catastrophic event. Although previous work has looked at identifying psycho-social risk factors for relapse at a population level ^(2;3;4;5)^, this is very different than predicting that a specific individual is at risk for relapse in the imminent future.

Regular toxicology screening and especially urine drug testing (UDT) is generally accepted as a part of outpatient medication for opioid use disorder therapy programs using buprenorphine or methadone. Results of drug screening are used to ensure that the patient is taking their prescribed medications and not using illegal or illicit substances.

This work explores whether results from laboratory testing can provide additional, non-obvious but useful information to the clinician in the care of these patients. Specifically, we wanted to explore whether machine learning methods applied to this data could provide predictive analytics useful in an outpatient treatment environment in predicting relapse in otherwise compliant patients.

## 2 Background

The most commonly used form of drug screening in opioid treatment programs uses samples of urine analyzed by the quantitative immunoassay (IA) method. This method is implemented on laboratory equipment found in most clinical laboratories and, with appropriate training and planning, can be provided cost effectively in addiction treatment centers.

Immunoassays use antibodies designed to bind with a specific drug (e.g. methadone), metabolite (e.g. the 6-MAM metabolite of heroin) or class of compounds (e.g. opiates) in a liquid sample. Immunoassay testing performed on a laboratory instrument provides a numeric value for the concentration of an analyte. This value is sometimes referred to as a “level”.

Immunoassays have varying degrees of sensitivity and specificity depending on the particular instrumentation, the antibodies used in the reagents, and the cutoff value used. A cutoff value is the level of analyte that needs to be detected in a sample for it to be considered positive. Test results are considered positive if there is enough drug or metabolite present in a specimen to react with a predetermined threshold of antibodies in the assay.

When using a cutoff, a negative result does not exclude the presence of a drug or metabolite in a specimen, but reflects it was not in sufficient amount to cross the cutoff limit. Screening tests will use cutoffs chosen to minimize the incidence of false positives. Consequently this, increases the incidence of false negatives. Many laboratories use screening cutoff levels calibrated for workplace or law enforcement drug testing. These cutoffs may be set high to identify individuals that use large amounts of a substance and minimizes false positives from accidental environmental exposure, e.g. from second-hand cannabis smoke.

Further, because certain antibodies bind to classes of drugs, care must be taken in the design of the testing panel, and when interpreting results in the context of determining whether a patient is compliant with therapy. Some laboratories provide confirmatory testing using mass spectrometry, as well as individual review of each patient’s test results by a trained expert. Although more accurate, the cost of this type of testing can be prohibitive for regular use in some addiction treatment centers including state funded health insurance programs.

A sample of a drug screen report is shown in Appendix 1. Reports can be provided in a hardcopy format, or as part of the patient’s electronic medical record. Some laboratories indicate only “pass/fail” or “positive/negative” on the report, although other laboratories may also list the concentration of each analyte along with the cutoff or threshold level. Addiction treatment centers with in-house testing typically link their laboratory equipment with their EHR system so that the results from testing are available to the clinician for scheduled meetings with patients.

(More details of the science behind drug testing in addiction medicine can be found in Appropriate Use of Drug Testing in Clinical Addiction Medicine, Copyright © 2017 American Society of Addiction Medicine, see Reference (6).)

## 3 Methods and Results

The hypothesis we explored in developing predictive analytics for addiction medicine was to use the data obtained from this laboratory testing, in conjunction with information from vital signs and basic demographics. We note that:

1. The laboratory testing results contain a large amount of quantitative data that is largely ignored.
2. The detailed interpretation of laboratory testing data requires knowledge of pharmacology, molecular biology, and the inner details of the working of complex testing equipment.
3. Most addiction medicine physicians are unlikely to have this knowledge, and thus rely on the simplified reports provided.
4. This data could be amenable to machine learning and data science methods now being applied in other areas of medicine ^(7; 8; 9; 10)^, being tempered with the knowledge of how the underlying testing works, and the insights into patient behavior provided by expert clinicians.

Our hypothesis was that a patient who was characterized as responding well to therapy would have laboratory results over time, and that significant changes or patterns of variation would be correlated with future of incidents of relapse. We specifically wanted to see whether changes over time in a patient’s quantitative immunoassay results were correlated with future incidents of relapse.

Our first data set consisted of de-identified data from 283 subjects participating in a hospital-based outpatient buprenorphine program. The de-identified data set was obtained from Diagnostic Laboratory Medicine (Bedford MA) for a single practice. The de-identified data set contained age and gender laboratory test results.

Immunoassay results with specific concentrations (levels) were available for alcohol, amphetamines, barbiturates, benzodiazepines, buprenorphine, cocaine, MDMA (ecstasy), ethyl glucuronide (alcohol metabolite), fentanyl, 6-acetylmorphine, methadone, opiates, oxycodone, THC, and tramadol. Each data point was normalized to the cut-off level of the analyte. A “relapse” was defined as a single opioid test of 300ng/ml or higher, which is how this program defined an opioid positive. (For comparison, the US Department of Transportation uses as a cut-off 2000ng/ml when drug testing transportation workers.)

We excluded subjects with less than four weeks of data, as well as subjects whose lab tests indicated they were not taking their prescribed buprenorphine. A subject whose first recorded drug screen was opiate positive would have that record excluded. For subjects with a positive opiate test that subsequently had at least four previous negative opiate tests, we split their data into compliant and non-compliant time periods and add both to the data pool. For all subjects we used a maximum of ten test results. Of the 283 subjects in our first data set, we examined a three-month period and identified 20 subjects with a relapse >300 ng/ml of opioids with at least four previous consecutive weeks without a positive opiate test. We then identified 119 subjects with at least four previous consecutive tests without a positive opiate test.

For each subject we calculated several statistical measures, including the maximum analyte concentration and the coefficient of variance for each analyte. We also attempted to construct combined measures for multiple analytes that play similar roles in clinical practice. For example, alcohol and cannabis are used recreationally, and some subjects claim to use alcohol and/or cannabis to mitigate the cravings that lead to opioid misuse. If this were indeed the case, we would expect to see a stable score to be indicative of non-relapsing subjects. Another example is that cocaine and ecstasy are both often used in communal settings, and that a person with subthreshold but meaningful levels may have contamination from other substances such as cannabis.

We then used machine learning based optimization techniques across primary and secondary measures to distinguish between relapsing and non-relapsing individuals. We first took a subset of subject data limited to the 20 relapsing subjects and 30 non-relapsing subjects selected at random and limited the number of primitives as inputs to the model. The scoring algorithm was able to correctly identify 13 of the 20 relapsing subjects (30% false negative rate) and did not flag any of the non-relapsing subjects. We then ran all remaining subjects through the classification algorithm; only 7 of the entire pool of 119 non-relapsing subjects were flagged (5.9% false positive rate). A confusion matrix and model performance metrics are given in Figure 1.

**Figure 1.**
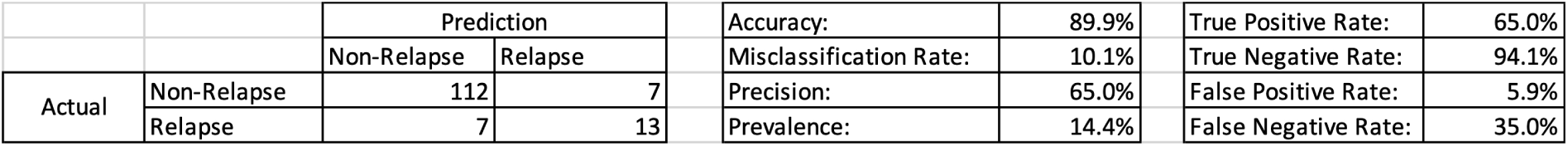
Confusion Matrix

We next looked at improvements to the predictive classifiers by adjusting and adding additional primitives, adjusting the look-back time period, and using different optimization techniques.

Since our goal is to produce a predictive measure of risk of relapse that can be used in clinical practice, we adjusted the algorithm to produce a risk score on a numeric scale that could be mapped to risk categories, so that a busy clinician treating these patients could easily understand the clinical significance of these classifiers to make treatment decisions in real time. We called this an “APRIL” score, an anacronym for “Analytic Prediction of Relapse Using Immunoassay Levels”. The APRIL score is defined for each patient on a linear 0-100 numeric scale, and is predictive of relapse in the following week.

Visual inspection of the resulting scores showed a clear demarcation of categories (Table 1). Only three relapsing subjects had scores between 30-100. Ten of the twenty relapsing subjects, and seven of the one-hundred nineteen non-relapsing subjects had scores between 10-30. The remaining seven relapsing subjects and one-hundred twelve non-relapsing subjects between 0-10. Conceptually this easily maps into a familiar risk category system of “red/yellow/green light”, with interventions to be targeted at the red and yellow categories.

**Table 1.** Subjects as Scored by Risk Categories

| Risk Category | APRIL Score Range | Number of Predicted Relapsing Patients | Number of Predicted Non-Relapsing Patients |
| --- | --- | --- | --- |
| RED | 30-100 | 3 | 0 |
| YELLOW | 10-30 | 10 | 7 |
| GREEN | 0-10 | 7 | 112 |

## 4 Discussion

In our APRIL score, the most weighted variables were the mean and variance of subthreshold levels of opiates, where the weights were computed by our machine learning algorithm from the training data. This is not surprising, as intuitively subjects that are still “using” at low levels are subjects at risk of relapse.

Surprisingly to us, on a linear scale of 0-100 with scores above 10 indicating risk of relapse, the average of all 13 relapsing subjects in both the yellow risk and the high red risk category had an average score of only 20. So simply noting the subthreshold levels of analytes on a traditional lab report would not easily identify subjects at risk.

Longer periods of baseline data improve performance. When we limited the data to only a maximum of four weeks of prior data, the false negative rate increased from 30% to 40% (6 -> 8 subjects out of 20), and the false positive rate increased from 5.9% to 9.2% (7 -> 11 subjects out of 119). Also, the most recent data does not appear to over-dominate the scoring. When we removed the most recent week (prior to relapse or non-relapse) of data from analysis, the false negative rate increased from 30% to 35% (6 -> 7 subjects out of 20), and the false positive rate increased from 5.9% to 7.6% (7 -> 9 subjects out of 119).

We also looked at how correlated alcohol and cannabis use are for risk of relapse. Measures of alcohol are correlated with 29.4% of subjects with an opioid relapse, and 8.4% of subjects without a relapse. Measures of THC are correlated with 64.7% of subjects with an opioid relapse, and 34.5% of subjects without a relapse. Although we would never suggest using only alcohol and cannabis use data to predict risk of opioid relapse, we do note that significant alcohol use (accuracy 82.7%, precision 37.5%) is more predictive of relapse than cannabis use (accuracy 65.5%, precision 24.1%).

## 5 Conclusions and Next Steps

We have developed an algorithmic method to identify patients in an opioid treatment program at high risk of imminent relapse.

Importantly, this method only uses data that is already being collected and included in a patient’s medical records, can be made available to the treating clinician before or during a patient visit, and by using a red-yellow-green risk categorization can be presented in an easy to understand way. With this a clinician can perform a targeted intervention to avert a likely relapse event in the week before it occurs.

Proactively identifying two-thirds of otherwise compliant patients that had a serious relapse represents a significant advance over the state of the art. The misidentification of patients as high-risk is low (<6%), so clinicians can have confidence that targeted interventions are worthwhile.

Further work with expanded data sets including larger patient populations and information in electronic health records may increase the clinical utility of this method. Also, the use of behavioral indicators, i.e. data available about decisions a person makes outside of the medical system, may allow us to expand the predictive power to identify patients with long periods of sobriety but who are also at high risk. This use of expanded data sets is central to the development of dynamic scoring systems that would allow the personalization of the predictive algorithms for each patient based on their on-going course of care. (See Figure 2.)

**Figure 2.**
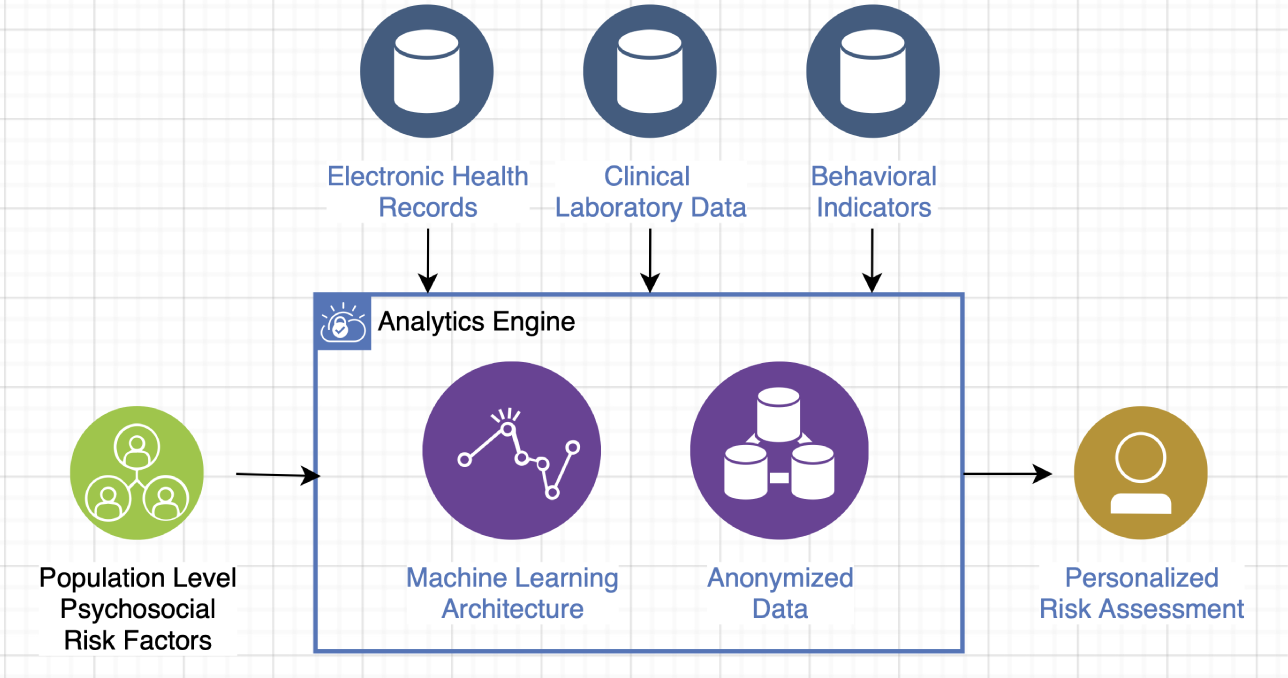
Architecture for Dynamically Scored Risk Assessment

We look forward to testing the use of these methods in clinical practice, to see if targeted interventions in patients identified as at risk makes a meaningful impact in reducing the actual rate of relapse.

## Data Availability

Data in not available for third party use.

## Appendix 1. Sample Urine Drug Testing Report

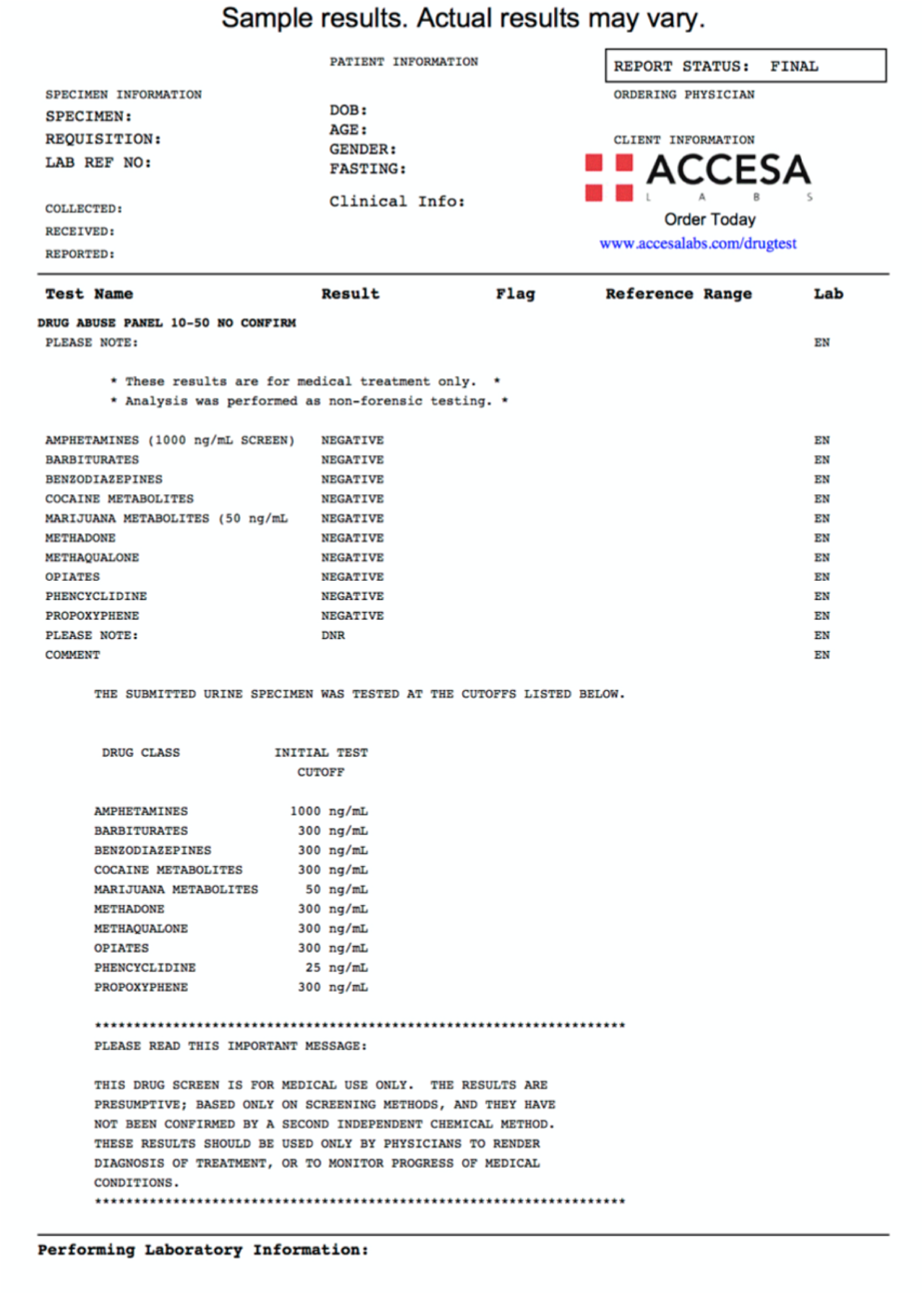

## Author biography

**Paul Pyzowski** Paul Pyzowski is CEO of Altimate Health, a company focused on innovation by applying data science to behavioral health. He previously held senior management positions in neuromodulation companies developing novel therapies for treatment of Parkinson’s disease and pediatric epilepsy. He managed a specialty clinical laboratory providing services to substance use disorder programs in hospitals and out-patient settings in New England. Mr. Pyzowski obtained his BS in Electrical Engineering from Carnegie-Mellon University where he studied computational physics, and spent the first decade of his career in applied numerical methods for computational design.

**Barbara Herbert** Dr. Barbara Herbert is a Fellow of the American Society of Addiction Medicine (ASAM), Diplomat of the American Board of Addiction Medicine, and immediate past president of the Massachusetts Chapter of ASAM. She was a member of the first advisory committee of the Massachusetts Physicians Monitoring Program, and the Governor’s Task Force to Address Addiction. Dr. Herbert was chief of addiction medicine for the fourteen hospital Steward Healthcare System including St. Elizabeth’s Comprehensive Addiction Program (SECAP), Massachusetts’ oldest and most established center focused on the treatment of substance use disorders. She has served in clinical and management roles at Commonwealth Care Alliance and Column Health.S he graduated from State University of New York at Stony Brook, School of Medicine and completed her residency at the Hospital of the University of Pennsylvania and a post doc fellowship at Harvard University.

**Wasim Q Malik** Dr. Wasim Q. Malik is an Assistant Professor in the Department of Anesthesia, Critical Care and Pain Medicine at Massachusetts General Hospital, Hospital Medical School, where he is the Director of the Neuromotor Signal Processing Laboratory. He serves on the Steering Committee of the IEEE Brain Initiative, on the Business Advisory Board of the Epilepsy Foundation, and as the Chair of the IEEE Engineering in Medicine and Biology Society’s Boston Chapter. He received his DPhil in electrical engineering from Oxford University and postdoctoral training in computational neuroscience from MIT.

